# Genomic characterization and Epidemiology of an emerging SARS-CoV-2 variant in Delhi, India

**DOI:** 10.1101/2021.06.02.21258076

**Authors:** Mahesh S Dhar, Robin Marwal, VS Radhakrishnan, Kalaiarasan Ponnusamy, Bani Jolly, Rahul C. Bhoyar, Viren Sardana, Salwa Naushin, Mercy Rophina, Thomas A Mellan, Swapnil Mishra, Charles Whittaker, Saman Fatihi, Meena Datta, Priyanka Singh, Uma Sharma, Rajat Ujjainiya, Nitin Batheja, Mohit Kumar Divakar, Manoj K Singh, Mohamed Imran, Vigneshwar Senthivel, Ranjeet Maurya, Neha Jha, Priyanka Mehta, A Vivekanand, Pooja Sharma, VR Arvinden, Urmila Chaudhary, Namita Soni, Lipi Thukral, Seth Flaxman, Samir Bhatt, Rajesh Pandey, Debasis Dash, Mohammed Faruq, Hemlata Lall, Hema Gogia, Preeti Madan, Sanket Kulkarni, Himanshu Chauhan, Shantanu Sengupta, Sandhya Kabra, The Indian SARS-CoV-2 Genomics Consortium (INSACOG), Ravindra K. Gupta, Sujeet K Singh, Anurag Agrawal, Partha Rakshit

**Affiliations:** National Centre for Disease Control, Delhi, India; CSIR-Institute of Genomics and Integrative Biology, New Delhi, 110007, India; Academy for Scientific and Innovative Research, Ghaziabad, 201002, India; Medical Research Council (MRC) Centre for Global Infectious Disease Analysis, Jameel Institute, School of Public Health, Imperial College London, UK; Department of Mathematics, Imperial College London, London, UK; Section of Epidemiology, Department of Public Health, University of Copenhagen, Denmark; Department of Medicine, Cambridge Institute of Therapeutic Immunology & Infectious Disease (CITIID), Cambridge, UK; Africa Health Research Institute, KwaZulu-Natal, South Africa

## Abstract

Delhi, the national capital of India, has experienced multiple SARS-CoV-2 outbreaks in 2020 and reached a population seropositivity of over 50% by 2021. During April 2021, the city became overwhelmed by COVID-19 cases and fatalities, as a new variant B.1.617.2 (Delta) replaced B.1.1.7 (Alpha). A Bayesian model explains the growth advantage of Delta through a combination of increased transmissibility and partial reduction of immunity elicited by prior infection (median estimates; ×1.5-fold, 20% reduction). Seropositivity of an employee and family cohort increased from 42% to 86% between March and July 2021, with 27% reinfections, as judged by increased antibody concentration after previous decline. The likely high transmissibility and partial evasion of immunity by the Delta variant contributed to an overwhelming surge in Delhi.

**One-Sentence Summary:** Delhi experienced an overwhelming surge of COVID-19 cases and fatalities peaking in May 2021 as the highly transmissible and immune evasive Delta variant replaced the Alpha variant.

## Introduction

After escaping relatively unscathed during the first wave of the COVID-19 pandemic, India witnessed a ferocious second COVID-19 wave, starting in March 2021 and accounting for about half of global cases by the first week of May. SARS-CoV-2 had spread widely throughout India in the first wave, with the third national serosurvey in January 2021 finding that 21.4% of adults and 25.3% of 10 -17 year old children were seropositive (1). Delhi, the national capital, was not included in the national serosurvey but had undergone multiple periods of high transmission in 2020 (Fig 1A). In a district-wise stratified serosurvey conducted by the Delhi Government in January 2021, overall seropositivity was reported to be 56.13 % (95% CI, 55.49 – 56.77%), ranging from 49.09% to 62.18% across eleven districts (2). This was expected to confer some protection from future outbreaks.

**Fig. 1.**
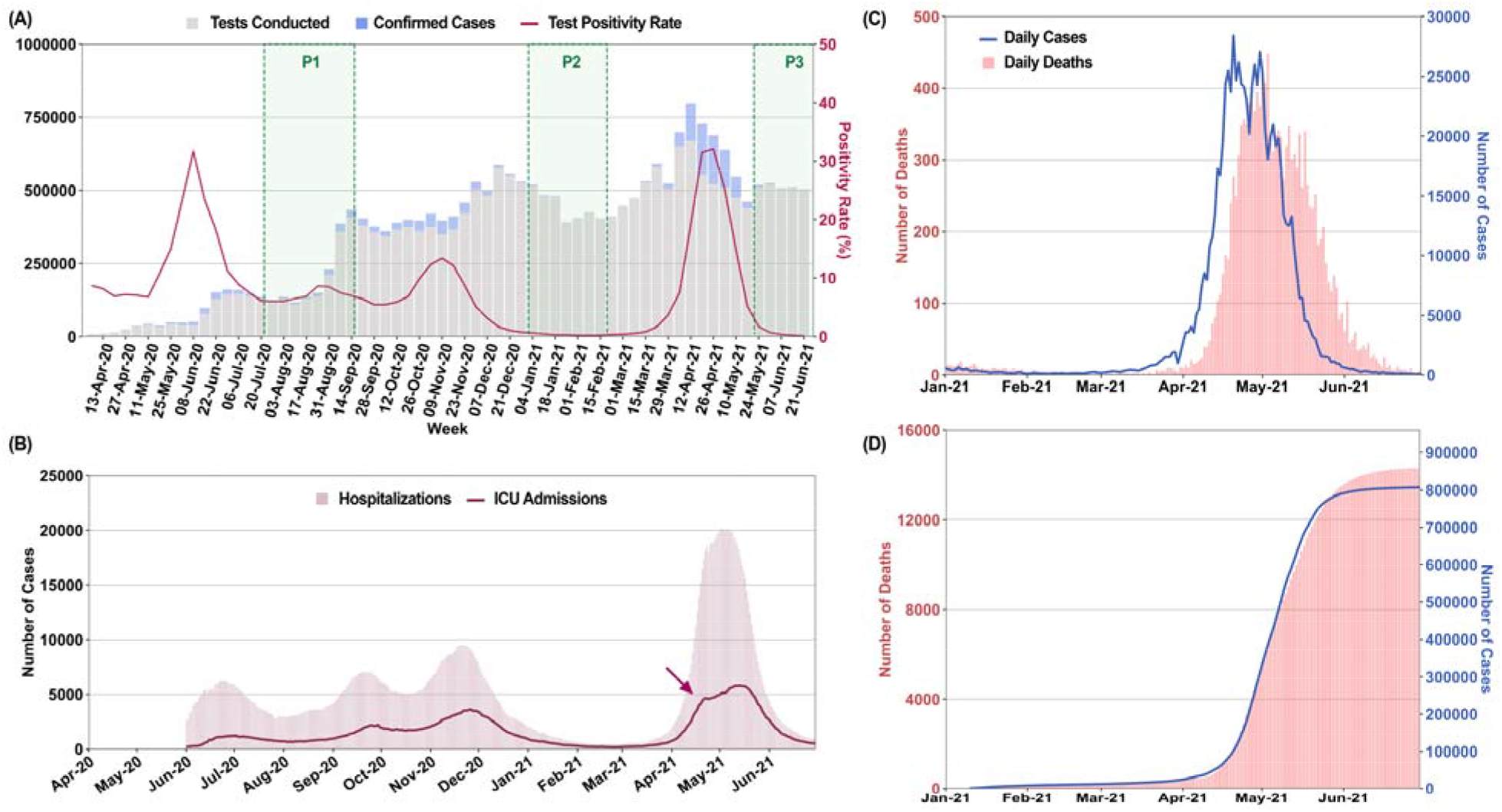
Multiple surges of SARS-CoV2 infections in Delhi with an overwhelming outbreak in April-May 2021. A) Weekly tests, new cases and test positivity rates (TPR) in Delhi from April 2020 to June 2021. Sample collection period for CSIR serosurveys is marked as P1-P3. B) Number of hospitalized and ICU patients plotted on a daily basis from June 2020 to 2021. Arrowhead marks possible saturation of ICU capacity (3) C) Daily cases and deaths from January to June 2021. D) Time advanced and scaled cumulative cases, fitted to cumulative deaths. Time advancement of cumulative reported cases by 8 days was done for maximal coincidence with scaled cumulative deaths. CFR = averaged scaling factor [cumulative deaths/time advanced cumulative cases]; (Mean +/- SD; 0.019 +/- 0.003).

Despite high seropositivity, Delhi was amongst the most affected cities during the second wave. The rise in new cases was exceptionally rapid in April, going from approximately 2000 to 20000 between March 31 and April 16. This was accompanied by a rapid rise in hospitalizations and ICU admissions (Fig 1B). In this emergency situation with saturated bed occupancy by April 12, major private hospitals were declared by the state as full COVID care-only and senior medical students, including from alternative medicine branches, were pressed into service (3). Deaths rose proportionately (Fig 1C) and the case-fatality ratio (CFR), estimated as the scaling factor between time-advanced cases and deaths (Fig 1D), was stable (mean, SD; 1.9, 0.3%). Population spread of SARS-CoV-2 is underestimated by test positive cases alone (1,2). To better understand the degree of spread and the factors leading to the unexpectedly severe outbreak, we used all available data including testing, sequencing, serosurveys, and serially followed cohorts.

In the absence of finely resolved or serial data from national and state surveys, we focussed on data for Delhi participants of a national serosurvey of Council of Scientific and Industrial Research (CSIR, India) employees and their family members (Fig 2A, table S1). Samples were initially collected from the end of July to mid-September 2020 (Phase I). Subsequently 2^nd^ and 3^rd^ surveys were held in January/February (Phase II) and end of May to early July 2021 (Phase III), bracketing the time period of interest. The cohort details and serosurvey methodology have been previously published (4).

**Fig. 2:**
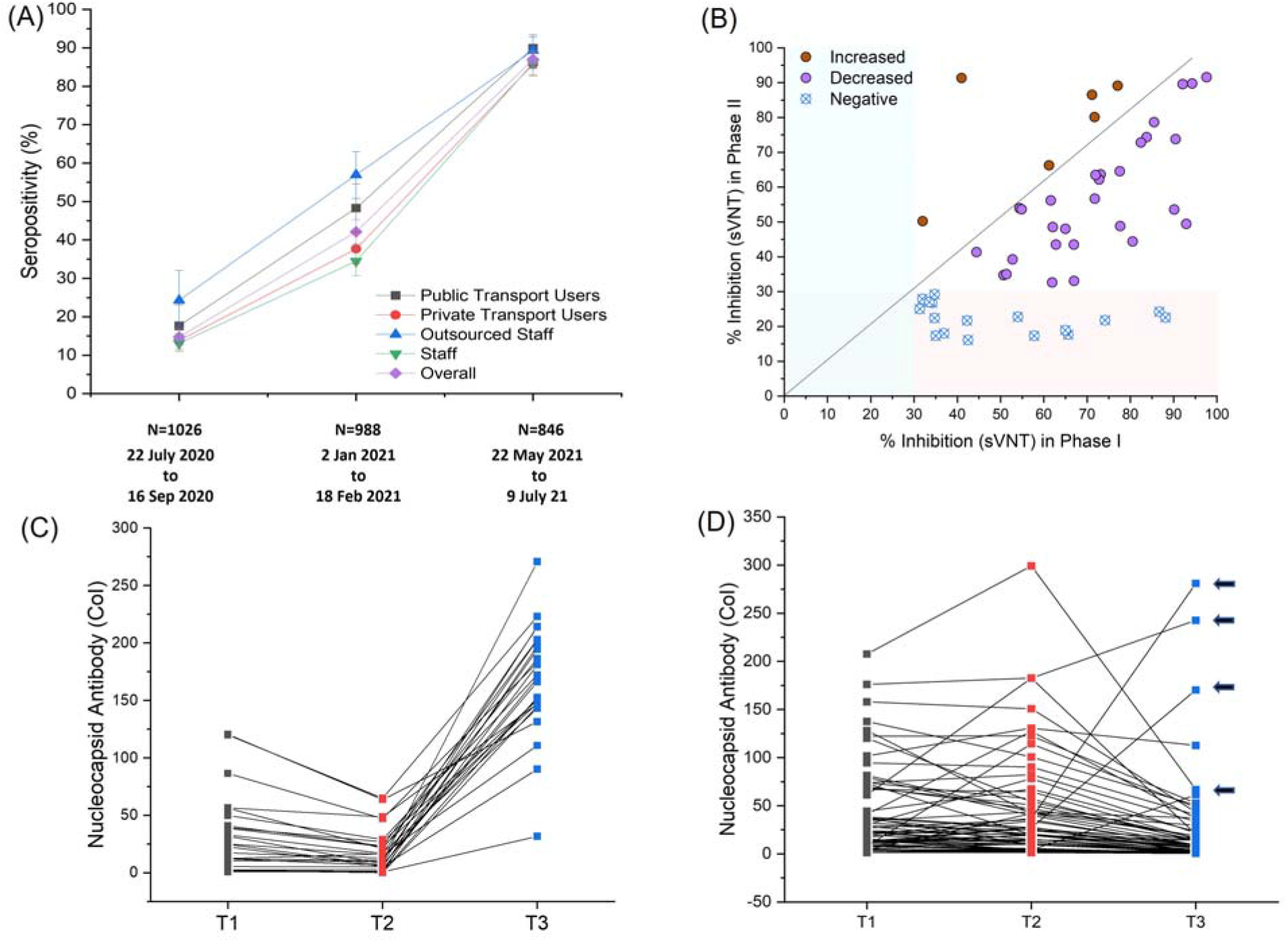
Serological estimates of prior infections, pre-existing immunity and new infections for the April-May outbreak. A) Seropositivity in CSIR cohort, sub-divided by nature of employment and use of public transport, is plotted for different time-periods (Phase I to Phase III, proportion +/- 95% CI). Details are in table S1. B) Variability and temporal decline in neutralization capacity estimated by sVNT assay between Phase I and II (n=52). C) Serial antibody concentration measurements in initially seropositive subjects (n=91). Pattern suggestive of reinfections is shown (decline followed by rise, n=25). D) shows remaining data (n=66), with four indeterminate reinfection cases indicated by arrowheads.

Infection was determined by anti-nucleocapsid assay, which is not affected by immunization with spike protein vaccines. The presence of neutralizing antibodies (NAb) to wild-type SARS-CoV-2 spike protein was estimated by a surrogate viral neutralization test (sVNT, Genscript). Previous results from the full cohort have been comparable to government serosurveys, but Delhi cohort values have been slightly lower. This may be due to an overrepresentation of members with the ability to reduce exposure and avoid public transport (Fig 2A). Within these limitations, the Delhi cohort showed a rise in seropositivity from 14.7% in Phase I (95% CI, 12.6-17.0%) to 42.1% in Phase II (95% CI, 39.0-45.2%). About one-third of NAb^+ve^ subjects at Phase I became Nab^-ve^ by Phase II, with most showing declining inhibition on sVNT assays (Fig 2B) (5). Phase III seropositivity increased to 87% (95% CI, 84.5-89.1%) amongst unvaccinated subjects. New infections between March and June 2021 are thus likely to have vastly exceeded known cases. Amongst ninety-one previously infected subjects with serial measurements at three time points including Phase III (T3), 25 (27.5%, 95% CI 18.4-37.5%) had a pattern of declining antibody concentration between T1 and T2, followed by a sharp rise at T3, indicative of reinfection (Fig 2C). Confirmation of reinfection by either RT-PCR (n=8) or symptomatic illness (n=2) was available for ten of the twenty-five subjects. No severe illness or hospitalization was reported in reinfections.

Time periods of increased transmission were associated with declining Ct values (Fig 3A, fig. S1), attributable to a higher proportion of recently infected individuals with high viral loads (6). However, the Ct decline was far greater in April 2021 (dCt, SE; -4.06, 0.27, p<0.001) than seen previously (dCT Nov 2020 ∼ -1.5). Comparing April 2021 with November 2020, high viral load samples (Ct<20) doubled in clinical samples (p<0.001), and nearly doubled in campus surveillance testing data, where most positives were from recently infected individuals (15% (n=297) vs 9% (n=358), p=0.02).

**Fig. 3.**
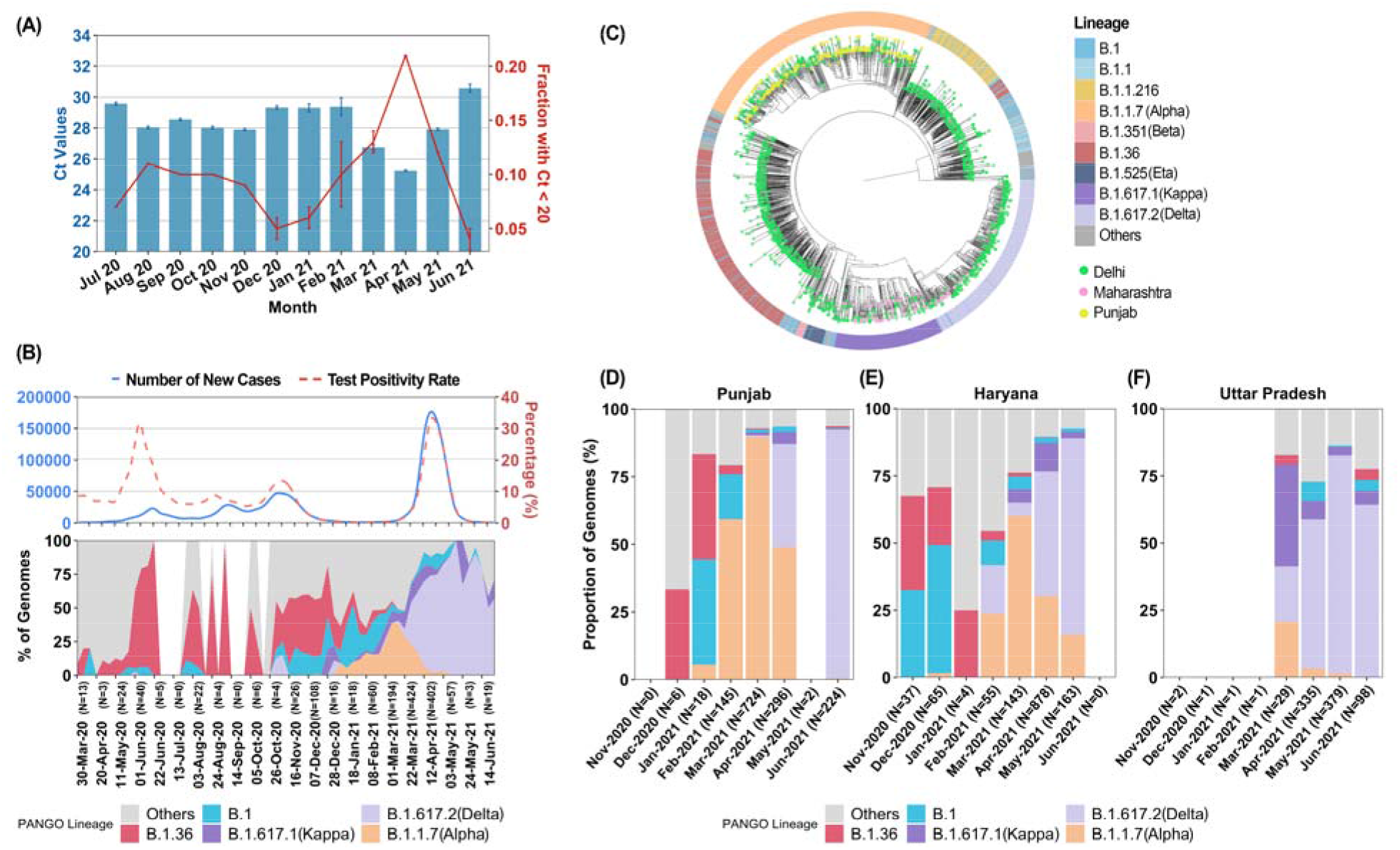
Genomic-Epidemiologic correlations. A) Time-trends of Ct values (mean +/- SE) and high viral load samples (proportion +/- SE) for Orf1 gene (E gene data, fig. S1). B) Smoothed graph of main lineages in Delhi from March 2020 to May 2021 in biweekly increments. New cases and TPR are aligned and plotted on the same timeline. C) Phylogenetic analysis for VOC strains between Delhi and states (Punjab and Maharashtra) with known VOC outbreaks before April 2021. Further analysis suggesting a super-spreading event for Alpha is shown in fig. S3. D-F) Month-wise proportions of different lineages (n>3) in states surrounding Delhi. Additional data is shown in fig. S4-S5.

Genome analysis trends, in representative samples drawn from general over the same period, showed seeding and expansion of B.1.1.7 (Alpha), B.1.617.1 (Kappa) and B.1.617.2 (Delta) lineages in 2021, with Delta becoming the dominant lineage in Delhi during April (Fig 3B). The proportion of Delta variant was strongly correlated to the rise in cases and healthcare stress (fig. S2). Overall, the genomic and epidemiological data were most consistent with the hypothesis that a new variant with higher infectivity, Delta, was driving the unexpected overwhelming surge in Delhi. Recent *in vitro* data supports the possibility of a higher replication rate for Delta, thereby explaining potentially higher viral loads in rtPCR data and greater transmissibility (7).

We further investigated the sequence of seeding and spread of Alpha, Kappa, and Delta variants of SARS-CoV-2. Phylogenetic analysis showed common origins between Alpha variants in Delhi and Punjab, and between Kappa or Delta variants in Delhi and Maharashtra, where Kappa and Delta were first sequenced (Fig 3C, fig. S3). There was a recurring pattern of initial smaller outbreaks with Alpha variant, followed by larger outbreaks coinciding with Alpha to Delta transition across all neighbouring states (ig. S4). The substantial relative growth advantage of Delta was explored in terms of transmissibility and/or immune escape. The rise of Delta, but not other lineages, was temporally coincident with rise in TPR and new cases during the surge (fig. S5). While overall vaccination levels were only about 5% in Delhi, most healthcare workers had received one or two doses of ChAdOx1-nCov19 (Astra-Zeneca / Serum Institute, India) or BBV152 (Bharat Biotech, India) (8,9). We sequenced twenty-four breakthrough infections starting at least one week after the first dose, collected between 22 March and 28 April at NCDC. The ratio of Delta to non-Delta lineages was 850:1211 from 20 March to 30 April. In contrast, Delta to other lineage ratio was 13:3 in sixteen breakthroughs post one dose and 7:1 in eight breakthroughs post the 2^nd^ dose of vaccine. While the small sample size and lack of a formal control group preclude definitive analysis, estimated higher odds for Delta in vaccination breakthroughs (OR, 7.1; 95% CI, 2.4-20.9) corroborate other reports of reduced vaccine effectiveness against Delta (10).

To better characterise how the properties of Delta might differ from other SARS-CoV-2 lineages previously circulating in the city, we used a Bayesian model of SARS-CoV-2 transmission and mortality that simultaneously models the dynamics of two-categories of virus (“B.1.617.2” and “non-B.1.617.2”) (11), whilst also explicitly incorporating natural waning of immunity derived from prior infection, with the duration of immunity consistent with the results of recent longitudinal cohort studies (12,13). Details of the model and input data are in Supplementary Methods. Briefly, the model is fitted to COVID-19 mortality data, genomic sequence data presented here and from GISAID (with lineage classification carried out used Pangolin (14,15) and serological data presented alongside an additional longitudinal serosurvey carried out in the city in the period July-December 2020 (16). Substantial uncertainties remain as to the date of introduction of B.1.617.2 into Delhi and the degree of COVID-19 death under-ascertainment. We therefore explored a range of different scenarios in which we varied under-ascertainment (10%, 33%, 50%, and 66%) and introduction dates (15 January 2021, 31 January 2021, 14 February 2021 and 28 February 2021).

Using this framework for an introduction date of 14 February 2021 and mortality under-ascertainment of 50%, our results shown in Fig. 4 indicate that the Delta variant is ×1.3-1.7 (50% bCI) fold more transmissible than earlier/co-circulating SARS-CoV-2 lineages in Delhi, including the Alpha variant. Importantly, the model also indicates that the Delta variant can partially evade immunity elicited by prior infection, with prior infection providing only 50 to 90% (50% bCI) of the protection against infection with Delta variant that it provides against previous lineages. We note there is an inherent trade-off between transmissibility and immune escape, and that the worst-case scenario of both very high transmissibility and immune escape is rejected *a posteriori* by the data. Fig. 4A also highlights the nature of uncertainty in the exact level of immune escape and transmissibility increase, since these inferred characteristics of the Delta variant are collinear given the modelling framework used and data currently available. However, the findings are robust to variation in prior assumptions, including both changing under-ascertainment and the date of introduction (Supplementary Methods; Tables S2-3). The results are valid for a population where the majority of immunity arose from prior infection (rather than vaccination), which is true for Delhi. Based on median estimates of the model (Fig 4A) and high transmissibility of the background Alpha lineage (17), Delta should potentially be at least twice as transmissible as other lineages.

**Fig. 4.**
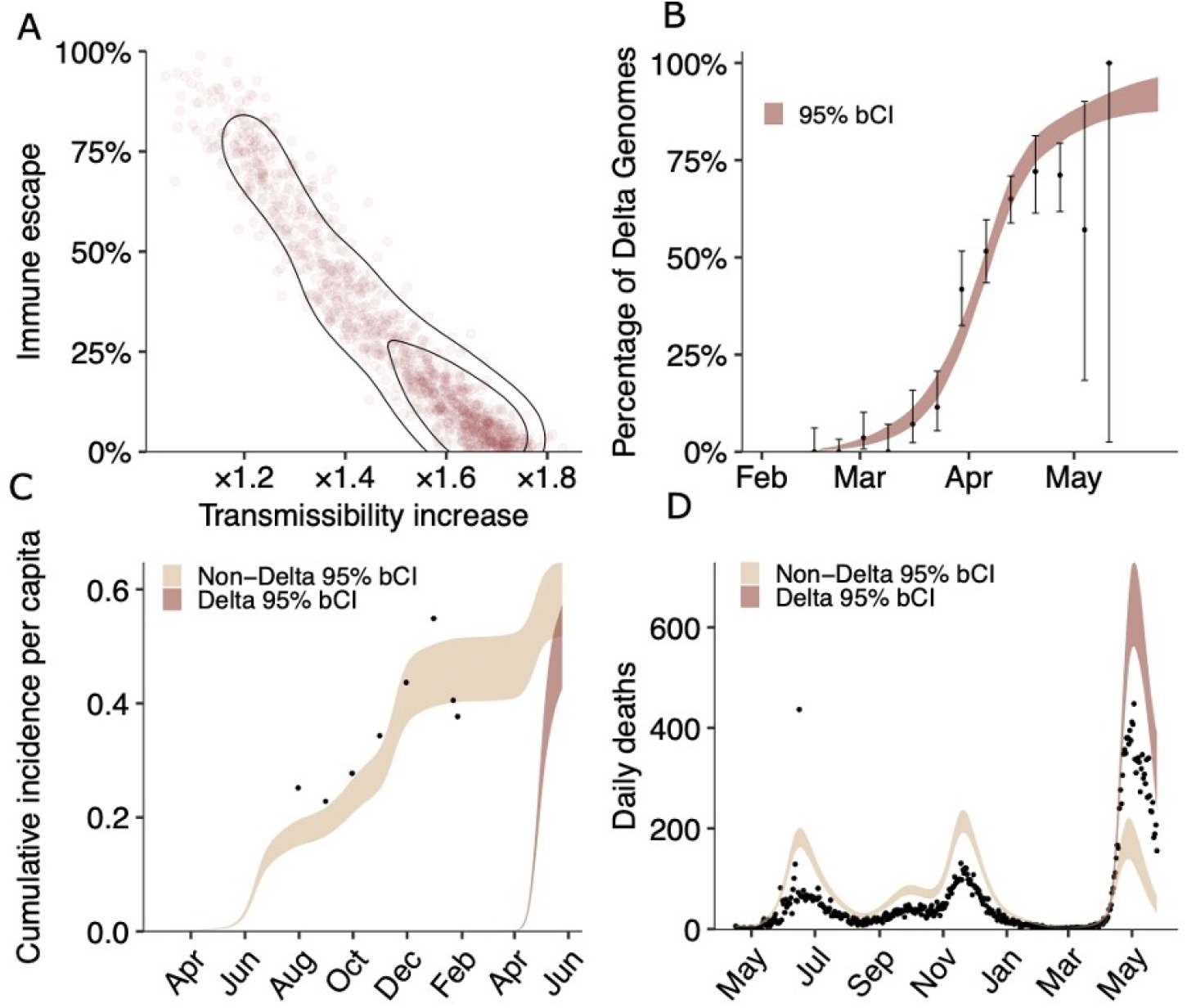
Estimates of the epidemiological characteristic of the Delta variant. Values were inferred from a two category Bayesian transmission model fitted to mortality, serosurvey and genomic data from Delhi, India. A. Joint posterior distribution of the Delta variant immune escape and transmissibility increase relative to non-Delta categories. Immune escape has a median of 20% with (10%-50%) bCI50%, and transmissibility increase has a median of ×1.5 with (1.3-1.7) bCI50%. B. Delta fraction over time inferred by the model. Black dots represent genome sampling data points, with exact binomial confidence intervals. C. Serosurvey data (black dots) and inferred cumulative incidence for Delta and non-Delta variant categories. D. Mortality data (black dots) and inferred deaths assuming 50% under reporting. Other under-ascertainment scenarios are presented in the Supplementary Information.

The transmissibility and immune escape properties of B.1.617.2 were unexpected since the immune escape associated Spike protein mutation E484Q was lost and transmissibility / immune escape associated Spike mutation L452R, previously reported in B.1.427 / B.1.429, was associated with much less severe outbreaks in California (18). A structural model representing B.1.617.2 spike protein provided plausible insights (fig. S6). We speculate that a combination of mutations at three critical sites in the Spike protein – N terminal domain deletions, Receptor Binding Domain (L452R, T478K), and S1/S2 cleavage site region (P681R, D950N) leads to an enhanced growth advantage of B.1.617.2 over other lineages by stabilizing the ACE2 receptor interaction and enhancing cleavage (see Supplementary Methods). Functional studies confirming the importance of these variants are reported elsewhere but a full understanding remains elusive (7,19).

Overall, we conclude that the Delta variant is capable of initiating fast-rising outbreaks and causing reinfections and vaccination breakthroughs. We emphasize that a strong public health response at the global level will be needed for its containment.

## Data Availability

Sequence data is being continuously uploaded on the GISAID.

https://www.gisaid.org/

## Acknowledgments

Suggestions by Prof Narendra K Arora, Prof G. Padmanabhan and member of the Scientific and Clinical Advisory Group of INSACOG are acknowledged.

This work is licensed under a Creative Commons Attribution 4.0 International (CC BY 4.0) license, which permits unrestricted use, distribution, and reproduction in any medium, provided the original work is properly cited. To view a copy of this license, visit https://creativecommons.org/licenses/by/4.0/. This license does not apply to figures/photos/artwork or other content included in the article that is credited to a third party; obtain authorization from the rights holder before using such material.

## Funding

Support from Ministry of Health and Family Welfare, Council of Scientific and Industrial Research (CSIR-Phenome India Grant, MLP-2007; InGen-CoV2, MLP-2005), and Department of Biotechnology (INSACOG), India, is acknowledged.

## Competing interests

Authors declare that they have no competing interests

## Data and materials availability

All data used in the analysis is accessible through supplementary materials as data files or repository links (https://github.com/banijolly/ncov-Delhi-Epidemiology) including accession numbers for all genomic data that has been deposited at GISAID. Code and data for replication of the Bayesian inference are available in the following repository: https://github.com/ImperialCollegeLondon/Delta_Variant_Delhi

## Supplementary Materials

Materials and Methods

Supplementary Text

Figs. S1 to S6

Tables S1 to S3

Supplementary data files and repository links

References 20-51

## Notes

### Competing Interest Statement

The authors have declared no competing interest.

### Funding Statement

Support from Ministry of Health and Family Welfare, Council of Scientific and Industrial Research, and Department of Biotechnology, India, is acknowledged.

### Author Declarations

The SARSCoV2 genomic study was approved by the NCDC Ethics Review Committee vide approval 2020/NERC/14. The serology study was approved by the Institutional Human Ethics Committee of CSIRIGIB vide approval CSIR IGIB/IHEC/2019_20

### Summary of Updates

Manuscript revised with better representation of data and one more co-author.

## References

1. M. V. Murhekar et al., SARS-CoV-2 seroprevalence among the general population and healthcare workers in India. International Journal of Infectious Diseases 108, 145–155 (2021).

2. Govt of National Capital Territory of Delhi (Twitter Feed of Health Minister. (2021), vol. 2021. https://twitter.com/satyendarjain/status/1356547817427226624?lang=en

3. Declaration of major private hospitals as COVID care only and engagement of 4th and 5th year MBBS students in govt hospitals GNCTD dt 12 April 2021. Orders F.23/Misc/COVID-19/DGHS/NHC/2020/ Pt XVIII/90-97 and No.01/H&FW/COVID-Cell/2020/07/ss4hfw/726. accessed at http://health.delhigovt.nic.in/wps/wcm/connect/doit_health/Health/Home/Covid19/Covid+19+Related+order+April+2021).

4. S. Naushin et al., Insights from a Pan India Sero-Epidemiological survey (Phenome-India Cohort) for SARS-CoV2. eLife 10, e66537 (2021).

5. J. M. Dan et al., Immunological memory to SARS-CoV-2 assessed for up to 8 months after infection. Science 371, eabf4063 (2021).

6. J. A. Hay et al., Estimating epidemiologic dynamics from cross-sectional viral load distributions. Science 373, eabh0635 (2021).

7. P. Mlcochova et al., SARS-CoV-2 B.1.617.2 Delta variant replication, sensitivity to neutralising antibodies and vaccine breakthrough. bioRxiv, 2021.2005.2008.443253 (2021).

8. Folegatti, Pedro M et al. Safety and immunogenicity of the ChAdOx1 nCoV-19 vaccine against SARS-CoV-2: a preliminary report of a phase 1/2, single-blind, randomised controlled trial. The Lancet 396, 467–478 (2020).

9. R. Ella et al., Efficacy, safety, and lot to lot immunogenicity of an inactivated SARS-CoV-2 vaccine (BBV152): a, double-blind, randomised, controlled phase 3 trial. bioRxiv, 2021.06.30.21259439 (2021)

10. J. Lopez Bernal et al., Effectiveness of Covid-19 Vaccines against the B.1.617.2 (Delta) Variant. New England Journal of Medicine 385, 585–594 (2021).

11. S. Flaxman, S. Mishra, A. Gandy, H. J. T. Unwin, T. A. Mellan, H. Coupland, C. Whittaker, H. Zhu, T. Berah, J. W. Eaton, others, Estimating the effects of non-pharmaceutical interventions on COVID-19 in europe. Nature. 584, 257–261 (2020).

12. C. H. Hansen, D. Michlmayr, S. M. Gubbels, K. Mølbak, S. Ethelberg, Assessment of protection against reinfection with SARS-CoV-2 among 4 million PCR-tested individuals in Denmark in 2020: a population-level observational study. The Lancet 397, 1204–1212 (2021).

13. V. J. Hall et al., SARS-CoV-2 infection rates of antibody-positive compared with antibody-negative health-care workers in England: a large, multicentre, prospective cohort study (SIREN). The Lancet 397, 1459–1469 (2021).

14. A. Rambaut et al., A dynamic nomenclature proposal for SARS-CoV-2 lineages to assist genomic epidemiology. Nature Microbiology 5, 1403–1407 (2020).

15. O’Toole Á, Scher E, Underwood A, et al. Assignment of Epidemiological Lineages in an Emerging Pandemic Using the Pangolin Tool. Virus Evolution, veab064 (2021).

16. A. Velumani et al., SARS-CoV-2 Seroprevalence in 12 Cities of India from July-December 2020. medRxiv, 2021.2003.2019.21253429 (2021).

17. N. G. Davies et al., Estimated transmissibility and impact of SARS-CoV-2 lineage B.1.1.7 in England. Science 372, eabg3055 (2021)

18. M. McCallum et al., SARS-CoV-2 immune evasion by the B.1.427/B.1.429 variant of concern. Science 373, 648–654 (2021).

19. Ferreira, Isabella et al. SARS-CoV-2 B.1.617 mutations L452 and E484Q are not synergistic for antibody evasion. The Journal of infectious diseases, jiab368.

