## Supplementary Methods and Figures for "Genomic characterization and Epidemiology of an emerging SARS-CoV-2 variant in Delhi, India"

**This PDF file includes:**

Materials and Methods
Supplementary Text
Figs. S1 to S6
Tables S1 to S3
References 20-51
Data Files S1 to S9 as separate files

### Materials and Methods

Ethics

All work conducted under this study was approved by Institutional Ethics Committees at NCDC and CSIR-IGIB. Testing and sequencing of positive samples for genomic surveillance is exempted from individual informed consent, being a mandated public health service of NCDC and CSIR-IGIB for public health purposes. The use of deidentified data generated through clinical and public health services for research was reviewed and approved under certificates CSIR-IGIB/IHEC/2020-21/01 Dt. 28.03.2020, and *NCDC Ethical review committee* No: 2020/NERC/14. The serosurvey was approved under certificate CSIR-IGIB/IHEC/2020-21/01 Dt. 28.03.2020 and CSIR-IGIB/IHEC/2020-21/02 Dt. 23.02.2021, for project entitled "Phenome India - A long-term longitudinal observational cohort study of health outcomes" with individual informed consent from each participant (4).

Sampling and Metadata Collection

Nasopharyngeal and throat swab samples from COVID-19 confirmed cases with Ct value < 25 were collected and transported to Biotechnology Division, National Centre for Disease Control (NCDC), New Delhi, from the various testing sites across different states in India as per the sampling strategy of Central Surveillance Unit (CSU) of Integrated Disease Surveillance Programme (IDSP), NCDC. 24 post-vaccination RT-PCR confirmed SARS-CoV-2 positive samples were also included in this study (Data S4). All patient details and metadata were filled on the patient identification form and were accompanied with the samples. A total of 11,335 samples were received for whole genome sequencing at NCDC between November 2020 to May 2021 which were processed for viral genome sequencing.

PCR Amplification, Viral Genome Sequencing and Assembly

Viral RNA was isolated from the patient samples using MagNA Pure RNA extraction system (Roche) following the manufacturer's instructions. Whole-genome sequencing of the viral isolates was done as per the COVIDSeq protocol by Illumina using the NextSeq 550 platform. A total of 376 samples per lot were processed in batches of 94 with indexes A-D by loading 1.4 pmol of the library on the 75 cycle High Output Kit flow cell. Approximately 20GB of data was generated by genome sequencing which was processed using the Illumina DRAGEN COVID Pipeline and DRAGEN COVID Lineage Tools (v3.5.1). The raw data sequencing data generated in binary base call format (BCL) from the NextSeq 550 instrument was demultiplexed to FASTQ files using bcl2fastq (Illumina, v2.20). The raw reads were aligned against the SARS-CoV-2 reference genome (NC_045512.2) following the pipeline (20). The minimum accepted alignment score was set to 12 and alignment results with scores <12 were discarded. The coverage threshold and virus detection threshold were set to 20 and 5 respectively. The variant calling target coverage which specifies the maximum number of reads with a start position overlapping any given position was set at 50. The consensus sequences generated for the samples at the end of the DRAGEN COVID Pipeline were used for downstream analysis including lineage assignment and phylogenetic analysis. Out of the 9,557 genome sequences generated, 7,858 sequences with complete metadata were used for further analysis.

Genome Datasets and Lineage Analysis

Two datasets were compiled to estimate the lineage frequency of SARS-CoV-2 for the state of Delhi and other surrounding states and union territories (UTs) in North India: Punjab, Haryana, Uttar

Pradesh, Chandigarh, Himachal Pradesh, Uttarakhand, Jammu and Kashmir, and Ladakh (Data S1). Dataset A comprises 7,858 genome sequencing data generated in this study. Dataset B comprises SARS-CoV-2 genome sequences from these states publicly available in GISAID (with collection dates up to 30th June 2021). Only those sequences from GISAID having complete date of collection (YYYY-MM-DD) were included in Dataset B and appended to Dataset A. For Dataset A, lineages were assigned to the genome sequences using the Pangolin tool (14,15) (version 3.1.8, pangoLEARN version 2021-07-28) to match the current version of lineage assignments on GISAID. The lineage data was segregated according to states and date of collection. State wise frequency of variant of concern B.1.617.2 (Delta) was plotted along with frequencies of B.1.617.1 (Kappa), B.1.1.7 (Alpha), B.1, B.1.36 and 'other variants'. For Delhi, proportions of the different lineages were calculated as weekly aggregates and for the period April 2020 to June 2021 and plotted in context with the weekly aggregates of the number of new cases of COVID-19 reported from the state and test positivity rate. The dataset of number of tests, confirmed cases and positivity rate for the state of Delhi was taken from the state level database maintained at NCDC (Data S2). For states other than Delhi, sequences with dates of collection between 1 November 2020 and 30 June 2021 were used to analyze lineage frequencies of the virus aggregated monthly. The dataset of the number of tests and confirmed cases for other states was accessed from https://covidtoday.github.io/backend/. A table describing the GISAID accession IDs, date of collection and lineages of the samples used to calculate lineage proportions in different states from Datasets A and B is given as Data S1. The acknowledgement table for genome sequences downloaded from GISAID is given as Data S3. All FASTA sequences included as Dataset A are available at https://github.com/banijolly/ncov-Delhi-Epidemiology. Details for the 24 post-vaccination samples and the respective genome sequence IDs are given in Data S4. Raw Ct values for genes ORF1a (Target 1) and E (Target 2) analysed for the time period between July 2020 – June 2021 is available as Data S5.

Phylogenetic Analysis

1787 genome sequences from the state of Delhi were used for phylogenetic analysis along with an additional 152 B.1.617.2 (Delta) and 271 B.1.1.7 (Alpha) genomes from the state of Maharashtra and Punjab respectively. The phylogenetic tree was constructed following the Nextstrain protocol for genetic epidemiology of SARS-CoV-2 (https://nextstrain.github.io/ncov/) (21,22). Briefly, the genome sequences were aligned against the SARS-CoV-2 reference genome MN908947 (20) using NextAlign (21) using the default parameters of the Nextstrain protocol. The first 100 and last 50 bases were masked in the resulting alignment file before further processing. A fast maximum likelihood phylogenetic tree was constructed using IQTREE2 (23) using a general time reversible model (GTR) by specifying a log-likelihood epsilon value of 0.05 for final model parameter estimation. The tree was further processed to resolve polytomies and rerooted to have sample hCoV-19/Wuhan/WH01/2019 (EPI_ISL_406798) as the root. The pipeline further processes the tree using TreeTime (22) to estimate a skyline coalescent using a fixed clock rate of $8x10^{-4}$. The resulting phylogenetic tree was visualized and annotated using the R package ggtree (24). The phylogenetic network of SARS-CoV-2 isolates from Punjab collected during February-March 2021 was visualized in Auspice (fig. S3). The tree files generated by the analysis are available at https://github.com/banijolly/ncov-Delhi-Epidemiology.

Serosurvey

The serosurvey was conducted through a voluntary participation wherein personnel working at CSIR labs/centers and their family members gave their blood samples in July-Sep 2020 (Phase 1) and January-February 2021 (Phase 2) and May-July 2021 (Phase 3) (details are given in table S1 and Data S6). The study was approved by the Institutional Human Ethics Committee of CSIR-IGIB vide approval CSIR-IGIB/IHEC/2019–20 & CSIR-IGIB/IHEC/2020-21/02 and carried out in over 40 CSIR laboratories and centers spread across the country. Blood samples (6 ml) were collected in EDTA vials from each participant and analyzed on site or transported to CSIR-IGIB, New Delhi for Analysis. Elecsys Anti-SARS- CoV-2 kit from Roche Diagnostics was used to detect antibodies to SARS-CoV-2 NucleoCapsid antigen. It is a qualitative kit which was used for screening and a Cut-off index COI >1 was considered seropositive. Positive samples were further tested for quantitative antibody titers using the same manufacturer's kit directed against the spike protein (S-antigen). An antibody levels >0.8 U/ml was considered sero-positive as per manufacturer's protocol. The detection range of this kit is from 0.4 U/ml to 250 U/ml. For samples, where values of >250 U/ml were obtained; appropriate dilutions were made. Neutralizing antibody (NAB) response directed against the spike protein (RBD site) was assessed using GENScript cPass kit which is a surrogatevirus neutralization test (sVNT). A value of 30% or above was considered to have neutralizing ability. sVNT neutralization assay data for Phase 1 and Phase 2 is available as Data S7 and data for subjects with and without reinfection is available as Data S8 and Data S9 respectively.

Protein annotation and modelling

SARS-CoV-2 genomes were annotated for amino-acid substitutions by SnpEff version 4.5. The annotation was done according to the SARS-CoV-2 reference genome (NC_045512) (20). The structural model of the spike in 1 RBD-up state was generated using cryoEM structure of the spike 1 RBD-up state (PDB ID: 6VSB) as a template (25). To generate ACE2 bound structure, we took the X-ray structure of human ACE2 bound to the RBD domain with PDB ID: 6M0J (26). Detailed modelling methodology is mentioned in our previous work (27). The structural mutant model of B.1.617.2 variant was generated using the structural model of ACE2-bound 1 RBD-up spike conformation as a reference. Each chain was mutated formissense mutations using ChimeraX (28) whereas deletions in each chain were introduced by employing Coot (29).

### Supplementary Text

Epidemiological Model

The model described here builds on a previously published model of SARS-CoV-2 transmission introduced in Flaxman et al, 2020 (30), subsequently extended into a two-category framework in Faria et al, 2020 (31). Replication code is available at https://github.com/ImperialCollegeLondon/Delta_Variant_Delhi.

**Model Specification** The model describes two categories, denoted $s \in \{1,2\}$. The population-unadjusted reproduction number for the first category is defined as

$$R_{s=1,t} = \mu_0 \, 2 \, \sigma(X_t) \,, \qquad 1$$

where $\mu_0$ is a scale parameter (3.3), $\sigma$ is a logistic function, and $X_t$ is an second-order autoregressive process with weekly time innovations, as specified in earlier work(32). The population-unadjusted reproduction number of the second category is modelled as

$$R_{s=2,t} = \rho \mathbf{1}_{[t_2,\infty)} R_{1,t} \,, \qquad 2$$

with

$$\rho \sim \text{Gamma}(5,5) \; \in [0,\infty) \,, \qquad 3$$

where $\rho$ is a parameter defining the relative transmissibility of category 2 compared to category 1 and $\mathbf{1}_{[t_2,\infty)}$ is an indicator function taking the value of 0 prior to $t_2$, and 1 thereafter, highlighting that category 2 does not contribute to the observed epidemic evolution before its emergence. The prior for $\rho$ is chosen because it weakly informative, setting 90% of prior mass a between $\times$ 0.4 and $\times$ 1.8 increase in transmissibility, while maintaining a neutral to conservative default in the context of increased transmissibility since it has a mean of $\times$ 1 and median of $\times$ 0.9.

Infections arise for each category according to a discrete renewal process (33,34)

$$i_{s,t} = \left(1 - \frac{n_{s,t}}{N}\right) R_{s,t} \sum_{\tau<t} i_{s,\tau} \, g_{t-\tau} \,, \qquad 4$$

where $N$ is the total population size, $n_{s,t}$ is the total extent of population immunity to category $s$ present at time $t$, and $g$ is the generation interval distribution.

The susceptible depletion term for category $s$ is modelled as

$$n_{s,t} = \sum_{\tau<t} i_{s,\tau} \, W_{t-\tau} + \beta_s (1 - \alpha_{s,t}) \sum_{\tau<t} i_{\backslash s,\tau} \, W_{t-\tau} \,. \qquad 5$$

where $\backslash s$ denotes not-$s$, under assumptions of symmetric cross-immunity with prior

$$\beta \sim \text{Beta}(2,1) \,. \qquad 6$$

Immune escape or the evasion of cross-immunity of Delta, as reported in the analysis, is defined as the complement of the cross-immunity parameter, that is $(1-\beta)$. The prior for $\beta$ has been chosen to reflect our default assumption that the mostly likely scenario is no evasion of cross-immunity. $W_{t-\tau}$ is the time-dependent waning of immunity elicited by previous infection, which is modelled as a Rayleigh survival-type function with Rayleigh parameter of sigma = 310, which produces 50% of individuals still immune after 1 year. The cross-immunity susceptible term $\alpha_{s,t}$ is modelled as

$$\alpha_{s,t} = \frac{(1-\beta_s)\sum_{\tau<t} i_{s,\tau} W_{t-\tau}}{N-\beta_s \sum_{\tau<t} i_{s,\tau} W_{t-\tau}} \,. \quad 7$$

Infections in Delhi are seeded for six days at the start of the epidemic from $t_1$ as

$$i_{1,t_1} \sim \text{Exponential}(1/\tau)\,, \quad 8$$

with

$$\tau \sim \text{Exponential}(0.03)\,, \quad 9$$

and the second category for six days from $t_2$, which is 14-02-2021 in the central scenario, as

$$i_{2,t_2} \sim \text{Normal}(1, 20^2) \in [1, \infty)\,. \quad 10$$

Non-unit seeding of the B.1.617.2 variant and the diffuse prior represent our uncertainty in the precise date and magnitude of B.1.617.2's introduction/importation into Delhi.

The model generates deaths via the following mechanistic relationship:

$$d_t = \sum_s \text{ifr}_s \sum_{\tau<t} i_{s,\tau}\, \pi_{t-\tau}\,. \quad 11$$

The infection fatality ratios of each of the categories ($\text{ifr}_s$) are given moderately informative priors:

$$\text{ifr}_s \sim \text{Normal}(0.25, 0.02^2) \in [0,100] \quad 12$$

with our central estimate based on the results of Brazeau et al (35) and adjusted for the demography of the city. We allow for variation around the estimate of 0.25 however, with the prior providing some support for IFRs in the range 0.15% - 0.35%. This range is similar to estimates by Banaji for Mumbai, a city with comparable demographics, for which an IFR is reported with 95% confidence intervals of (0.15%, 0.33%)(36). A limitation of the model is the assumption of homogeneous exposure across subsets of the population.

**Likelihood component 1.** The observation model uses three types of data from four sources. In the first, the likelihood for the expected deaths $D_t$, is modelled as negative-binomially distributed,

$$D_t \sim \text{NegativeBinomial}\left(d_t(1-\omega), d_t + \frac{d_t^2}{\phi}\right)\,, \quad 13$$

with mortality data $d_t$ and diffuse dispersion prior

$$\phi \sim \text{Normal}(0, 5^2) \in [0, \infty) . \quad 14$$

and underreporting factor $\omega$, which describes the degree of death underascertainment, e.g. a value of 0.25 means 25% of COVID-19 deaths are not reported (due, e.g. to limited testing). The central scenario chosen for death underreporting is 50%, due to the wide range values reported in available literature for Delhi and India(36–41). To mitigate the effect of the uncertainty in death underreporting, sensitivity tests are carried out a range of values, $\omega$ in {10%, 33%, 50%, 66%}, to ensure inferences are robust.

**Likelihood component 2.** The second likelihood is based on genomic data from individuals where infections were sequenced and where the sequence was uploaded to GISAID. Specifically, the proportion of sequenced genomes identified as B.1.617.2 at time $t$ are modelled with a binomial likelihood

$$G_t^+ \sim \text{Binomial}(G_t^+ + G_t^-, \theta_t) , \quad 15$$

with positive counts for B.1.617.2 denoted $G_t^+$ and counts for lineages not belonging to B.1.617.2 recorded as $G_t^-$. The success probability for B.1.617.2 positivity is modelled as the infection ratio

$$\theta_t = \frac{\tilde{\iota}_{2,t}}{\tilde{\iota}_{1,t} + \tilde{\iota}_{2,t}} , \quad 16$$

where $\tilde{\iota}_{s,t}$ is given by

$$\tilde{\iota}_{s,t} = \sum_{\tau \le t} i_{s,\tau} \, \kappa_{t-\tau} , \quad 17$$

to account for the time varying PCR positivity displayed over the natural course of a COVID-19 infection. The distribution $\kappa$ describes the probability of being PCR positive over time following infection, and is based on (42).

**Likelihood component 3.** Serological data are incorporated in our modelling framework, using results from the survey presented in this work, and from Velumani et al. (43). The observed seropositivity $(S_t)$ on a given day, $t$, is modelled as follows

$$S_t^+ \sim \text{Binomial}\left( S_t^+ + S_t^-, \nu_t \sum_{\tau \le t} i_{s,\tau} \, C_{t-\tau} \right) , \quad 18$$

where $C_{t-\tau}$ is the cumulative probability of an individual infected on day $\tau$ having seroconverted less seroreverted by time $t$. This distribution is empirical and based on (44). The term $\nu_t$ is a multiplicative random effect specified to mitigate likely biases in the serological data

$$\nu_t = 2\sigma(Y_t) , \qquad Y_t \sim \mu + \eta_t , \qquad \mu \sim N(0,1) , \qquad \eta_t \sim N(0, \delta) , \qquad \delta \sim N^+(0,1) , \quad 19$$

where $\sigma$ is the logistic function.

Eq *4* can be modified to account for population effects (decreasing susceptible population over time) such that no over-shooting happens due to discretization as follows (45,46):

$$i_{s,t} = (N - n_{s,t})\left(1 - \exp\left(-\frac{i_{s,t}}{N}\right)\right), \quad 20$$

The formula for $i_{s,t}$ is derived from a continuous time model on $[t-1, t]$. This is to avoid discrete time effects such as infections going above the total population $N$. Specifically, we assume that the infections $i(\Delta t)$ in $[t-1, t-1+\Delta t]$ are given by the differential equation $\partial i(\Delta t)/\,\partial \Delta t = i_t\left(1 - \left(n_{s,t} + i(\Delta t)\right)/N\right)$, which has the solution $i(1) = i_t$ as above.

**Modelling limitations** Inferences from the model are subject multiple limitations, arising from biases in the data, and in our choices of priors. In particular, the level of underreporting in Delhi is poorly characterised. This uncertainty is mitigated by sensitivity testing, as described in the Supplementary Information. Similarly, uncertainty in the temporal waning of immunity, the date of when Delta was first introduced to Delhi, and the IFRs of SARS-CoV-2 variants in Delhi, all provide sources of bias that we currently only mitigate through sensitivity testing. Furthermore, we note serological data is likely to be systematically biased. This is partially addressed through the inclusion of an additional term in the serology likelihood that provides a multiplicative random effect.

**Computational notes** The analysis uses R version 3.6.3. Inferences are based on 2000 iterations of Hamiltonian Monte Carlo using 2 chains, with rhat statistics confirmed to be less than 1.02. The inference is performed using rstan version 2.21.2. Replication code and data are available at https://github.com/ImperialCollegeLondon/Delta_Variant_Delhi.

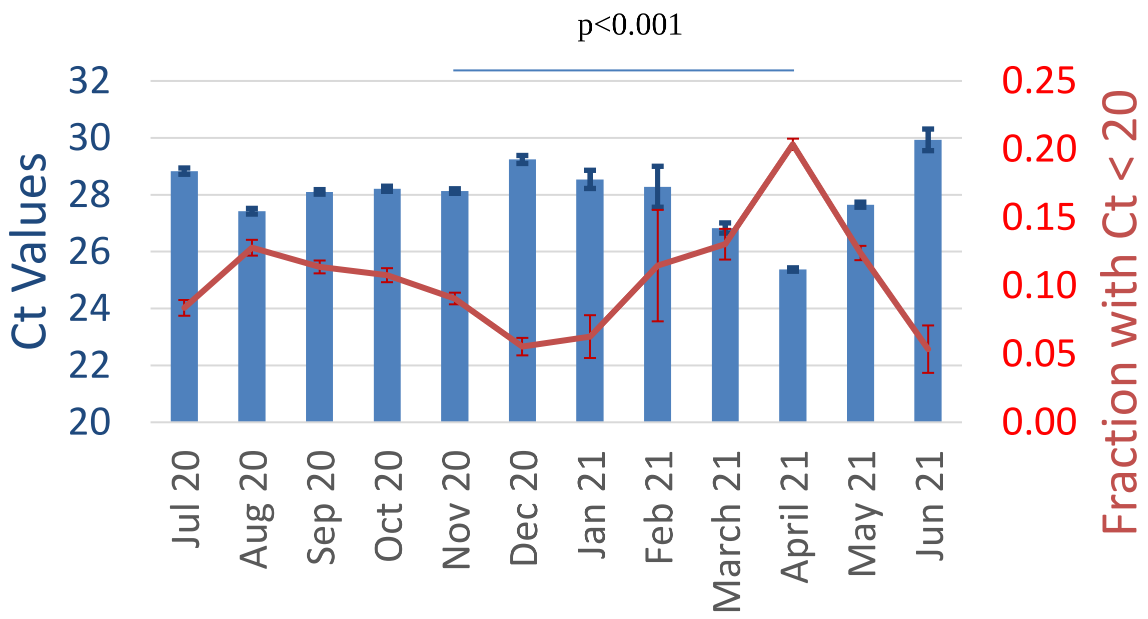


| Month / year | Positive samples (Ct <=35) |
|---|---|
| Jul 20 | 2303 |
| Aug 20 | 3332 |
| Sep 20 | 4552 |
| Oct 20 | 3666 |
| Nov 20 | 4555 |
| Dec 20 | 1281 |
| Jan 21 | 239 |
| Feb 21 | 61 |
| March 21 | 897 |
| April 21 | 9431 |
| May 21 | 3968 |
| Jun 21 | 168 |

**fig. S1. E gene Ct Values (blue) and Fraction of samples with Ct<20 (red) from July 2020 to June 2021.** Mean +/- SE (blue bar) is shown for Ct Values for monthly clinical samples testing positive on a single COBAS 6800 automated testing system at NCDC, Delhi. All Ct values less or equal to 35 were used for the analysis. Number of positive samples meeting the criteria is shown in the table. Proportion +/- SE is shown for fraction of high viral load samples (red line). Statistical significance of difference is calculated from two sample Z-tests for means and proportions between Nov 2020 and April 2021, as corresponding transmission surge time-periods. $p<0.001$

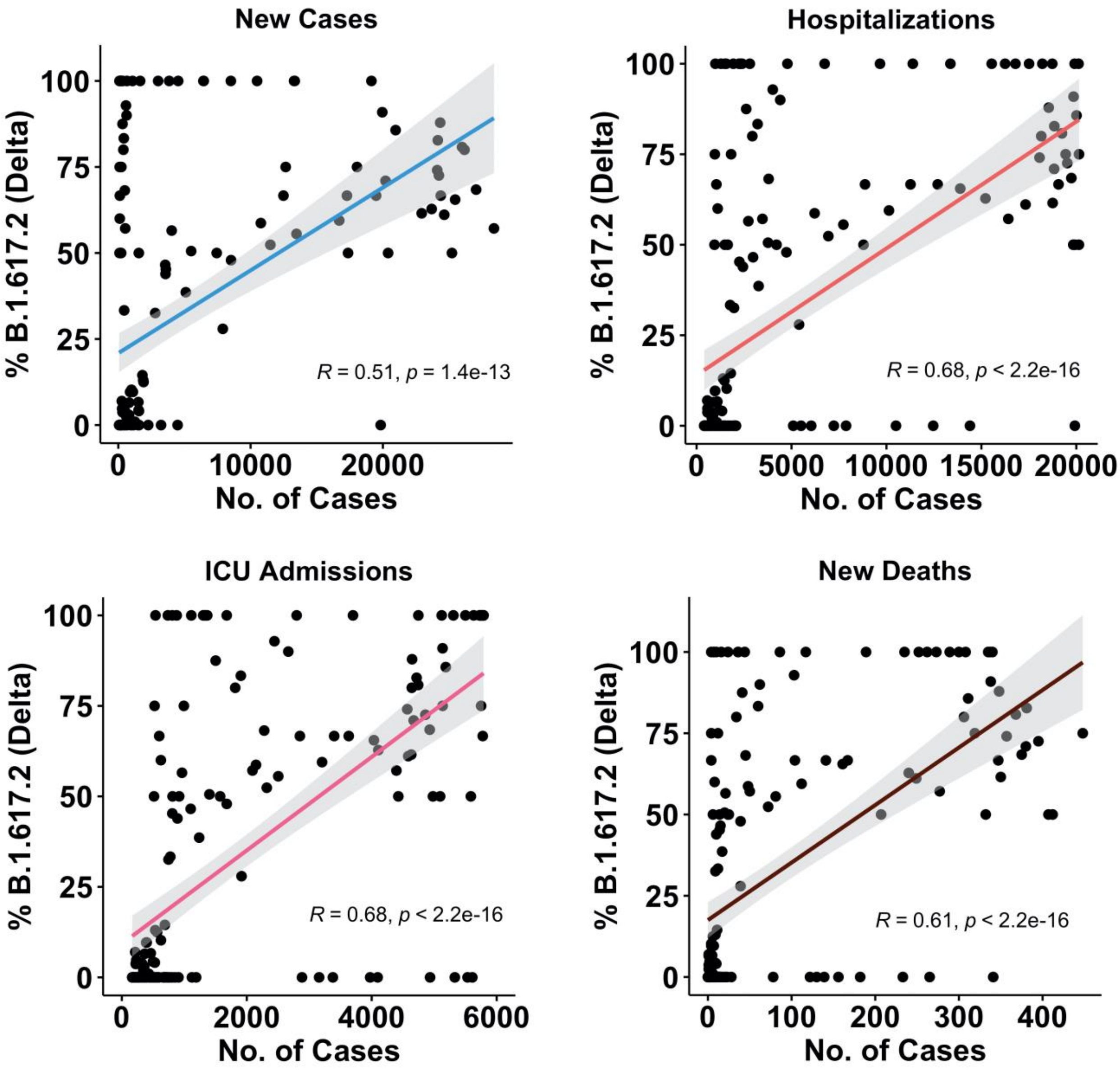


**fig. S2. Correlation plot between %Delta lineage (X-axis) and epidemiological variables for the April surge (Cases, Hospitalizations, ICU, Deaths).** Spearman coefficients were calculated for all epidemiological variables with % Delta.

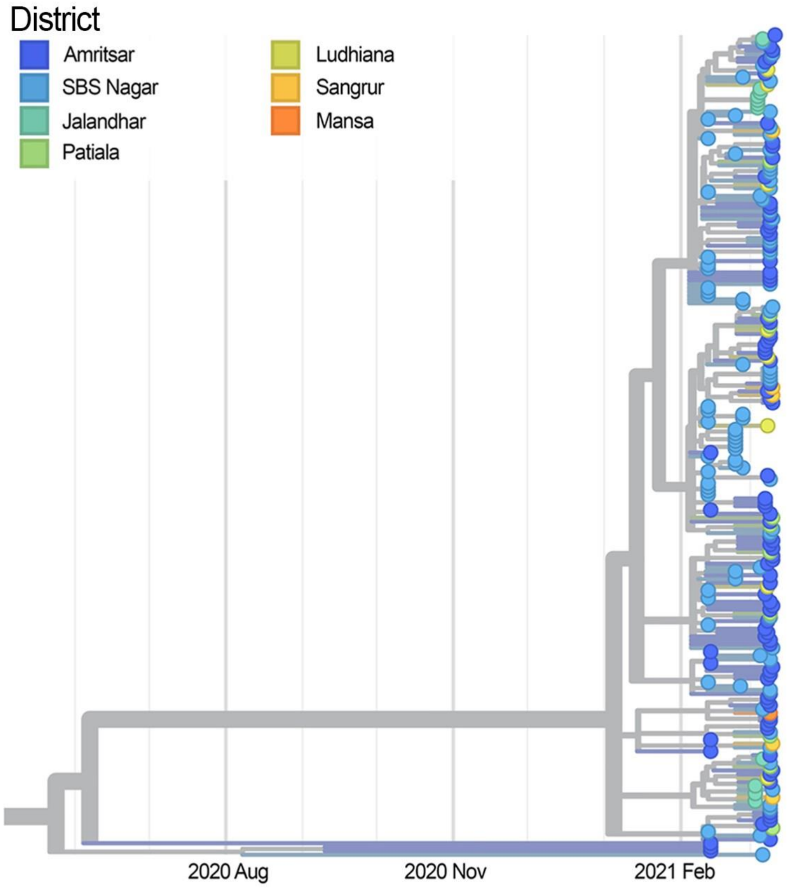


**fig. S3. Molecular signature of super-spreader event in Punjab.** Time-resolved phylogenetic tree for genome sequences from Punjab using samples collected during February-March 2021. Strong identity can be seen between sequences from various districts, corresponding to known social events during this period

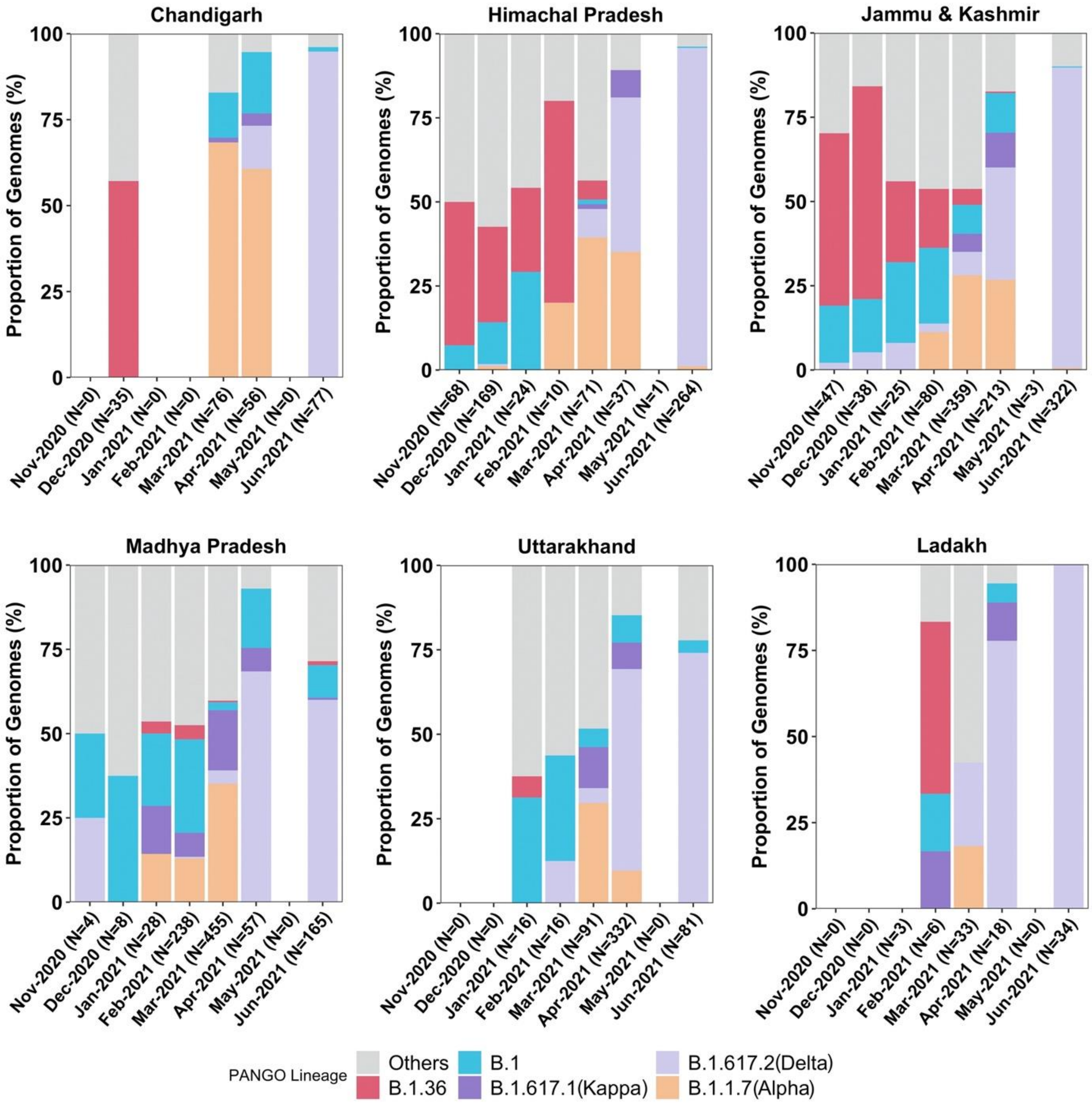


**fig. S4. Displacement of Alpha by Delta strain all over North India.** Normalized stacked bar graphs of main lineages for the states of Chandigarh, Himachal Pradesh, Madhya Pradesh, Uttarakhand, Jammu and Kashmir, and Ladakh. Outbreaks were seen in these states during April and May 2021, coincident with the rise of Delta (fig S5).

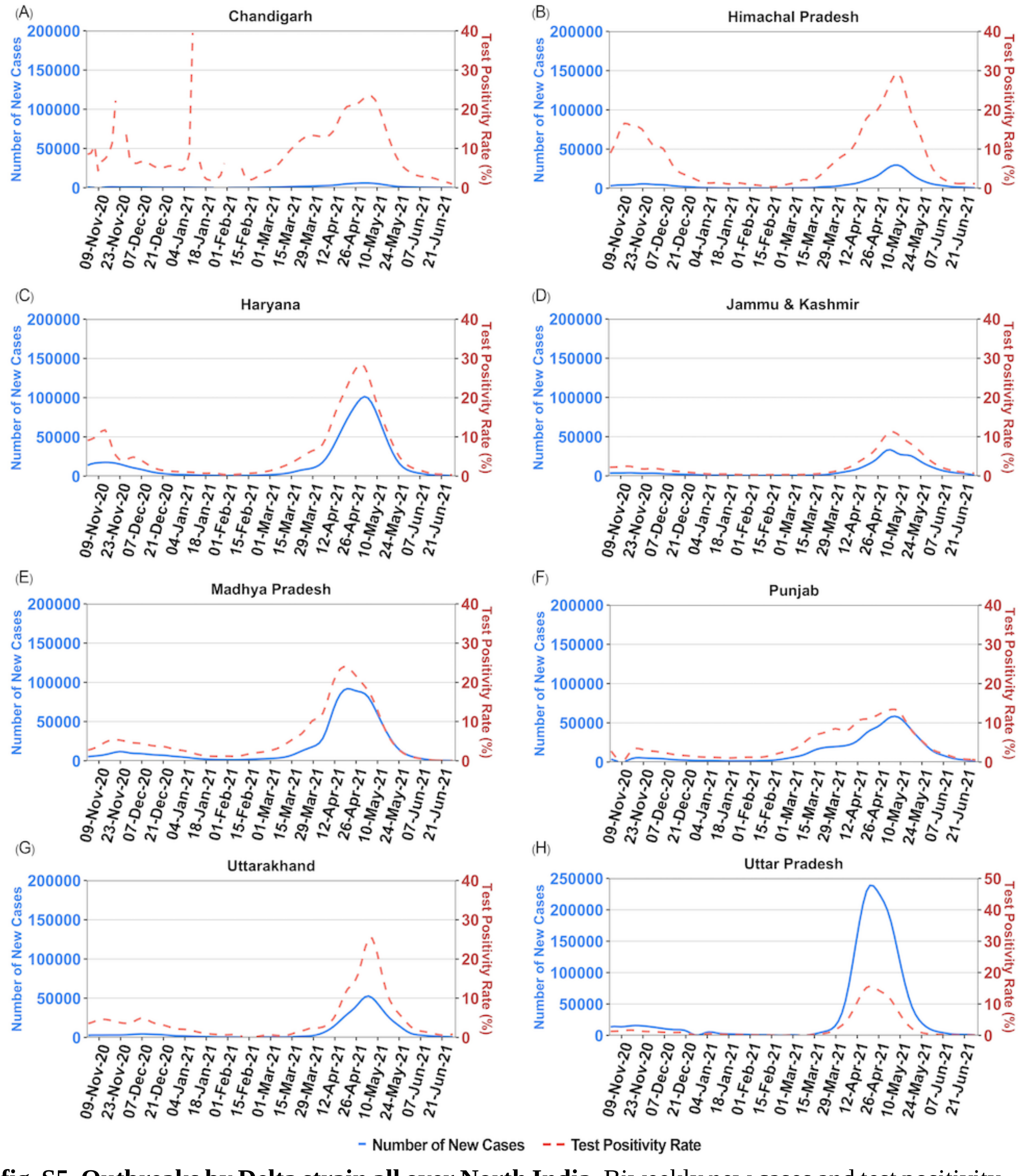


**fig. S5. Outbreaks by Delta strain all over North India.** Biweekly new cases and test positivity rates are shown for states around Delhi from November 2020 to June 2021 , coincident with the rise of Delta. Tests are a mixture of RT-PCR and antigen tests and separate positivity rates are not available. Peaks are coincident with Delta dominance on genome sequencing (Fig 3D-F and fig S4)

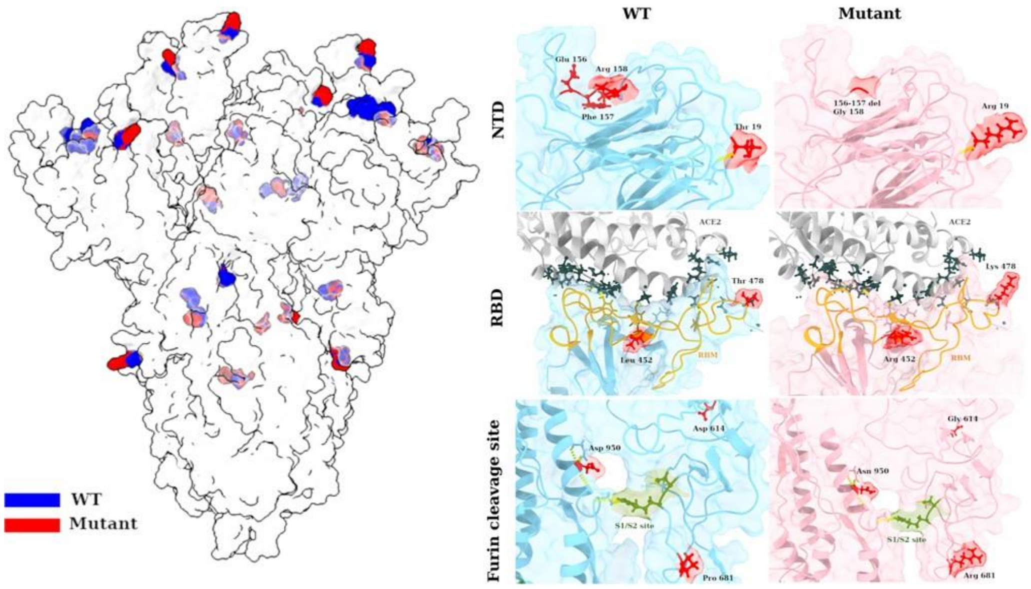


**fig S6. Mutant spike protein of B.1.617.2 lineage has critical mutations at furin cleavage and RBD sites that may enhance binding and cleavage.** The structural model of spike protein with seven mutations was generated and side-chains are highlighted in blue and red color to illustrate amino-acid substitutions. On the right panel, we show a zoomed snapshot of three critical regions namely, NTD, RBD, and furin cleavage sites. The resulting structural map provides insights into the plausible mechanisms of regulation of virus entry and binding. It contains seven mutations in the spike protein, excluding the predominant D614G substitution. Three of these mutations, two substitutions (T19R and R158G) and one deletion (ΔE156-F157), were found in NTD region. The six nucleotides spanning the entire stretch of deletion (ΔE156-F157) was juxtaposed with R158G mutation, with one nucleotide of glycine contributed from E156 and the rest two nucleotides contributed by R158, resulting in GGA codon for glycine. The mutations within NTD occur on N1 and N3 loops that composes the prominent mABs recognition sites (47,48). In our mutated model, R19 and G158 residues are surface accessible while in wild-type the T19, E156-F157 were relatively buried. We also found a unique RBD mutation T478K, in addition to previously reported L452R in B.1.617.2. Previous reports have associated L452R amino-acid change with antibody binding and has been classified as an escape variant (49,50). T478, on the other hand, is previously unidentified and is present directly on the receptor binding motif (RBM). The inherent long side-chain of mutated lysine reduces the gap with the ACE2 receptor as compared to the wild type. The distance between spike: K478 and ACE2:L85 is 8.3 Å, while the threonine maintains a distance of 10 Å. However, these distances may vary with other conformational states of spike RBD. We also observed significant increase in number of ACE2 residues around these mutated sites (L452R, T478K) in B.1.617.2. In contrast with 10 residues, 24 residues of ACE2 were in close proximity to the mutated side chains of B.1.617.2 RBD mutations. In addition, we observed two mutations (P681R and D950N) in proximity to the S1/S2 cleavage site. Previous reports have also shown that the dominant D614G, although distal to the furin cleavage site has an allosteric effect on conformational changes leading to RBD opening (51). The location of mutated residues is marked in red and, and critical regions such as RBM and S1/S2 site are highlighted for clarity.

**Table S1. Seropositivity in the Delhi CSIR cohort.** Seropositivity is shown for Phase I to III, along with 95% CI. In the beginning of the pandemic, with strong lockdowns, higher infection rates were seen in outsourced staff providing frontline and essential services, while laboratory employees and students were relatively protected. By mid-pandemic, with lockdowns lifted by July 2020, use of private vs public transport also determined risk. By the end of the Delta wave in Delhi, all subgroups reached similar seropositivity, suggesting universally high exposures. For comparisons between serially obtained values, or between sub-groups, statistical significance of difference of proportions was calculated via the standard 2x2 Chi-statistic; p<0.01 was considered significant.

| | Subjects | Seropositive (anti-NC antibodies) | Seropositive proportion overall (%) (95%CI) | Seropositive proportion (%)<br>Outsourced Staff vs Staff | Seropositive proportion (%)<br>Public vs Private transport users | Date of Collection | Age Range (Median) | M:F Ratio(%) |
|---|---|---|---|---|---|---|---|---|
| Phase I | 1026 | 151 | 14.72 (12.6-17.03) | 24.32 vs 13.11<br>p<0.01 | 17.65 vs 13.85<br>p=NS | 22 July 2020 to 16 Sep 2020 | 18-81 (39) | 74.7: 25.3 |
| Phase II | 988 | 416 | 42.11 (39-45.25)<br>p<0.001 vs Phase I | 56.93 vs 34.49<br>p<0.01 | 48.29 vs 37.67<br>p<0.01 | 2 Jan 2021 to 18 Feb 2021 | 18-84 (35) | 68.3:31.7 |
| Phase III | 846 | 736 | 87 (84.54-89.19)<br>P<0.001 vs Phase II and Phase I | 89.29 vs 86.05<br>p=NS | 89.96 vs 85.76<br>p=NS | 22 May 2021 to 9 July 21 | 18-82 (33) | 66.1:33.9 |

**Table S2.**

Inferred changes in epidemiological characteristics of B.1.617.2, depending on the timing of introduction assumed in the model, and level of under-ascertainment present in Delhi mortality data. Results presented are the median, with the 50% Bayesian Credible Interval, bCI, in brackets. Note that "Immune escape" refers specifically to the escape of immunity conferred by prior infection with other variants, rather than escape from immunity acquired through vaccination. It is further important to note that immune escape and transmissibility increase inferred for B.1.617.2 are values given with reference to the composition of earlier and co-circulating variants in Delhi, from the start of the epidemic to 25 May 2021.

| | | **Inferred epidemiological characteristic** | |
|---|---|---|---|
| **Timing of B.1.617.2 introduction** | **Mortality under-ascertainment** | **Immune escape** | **Transmissibility increase** |
| **15 Jan 2021** | 10% | 0.34 (0.16-0.60) | 1.48 (1.35-1.57) |
| **31 Jan 2021** | 10% | 0.39 (0.18-0.59) | 1.45 (1.36-1.55) |
| **14 Feb 2021** | 10% | 0.42 (0.21-0.64) | 1.47 (1.37-1.58) |
| **28 Feb 2021** | 10% | 0.45 (0.23-0.65) | 1.55 (1.45-1.65) |
| **15 Jan 2021** | 33% | 0.29 (0.11-0.55) | 1.48 (1.35-1.60) |
| **31 Jan 2021** | 33% | 0.29 (0.10-0.55) | 1.49 (1.35-1.59) |
| **14 Feb 2021** | 33% | 0.38 (0.16-0.64) | 1.47 (1.34-1.59) |
| **28 Feb 2021** | 33% | 0.43 (0.20-0.67) | 1.54 (1.42-1.67) |
| **15 Jan 2021** | 50% | 0.15 (0.07-0.35) | 1.54 (1.40-1.62) |
| **31 Jan 2021** | 50% | 0.15 (0.07-0.33) | 1.54 (1.40-1.61) |
| **14 Feb 2021** | 50% | 0.22 (0.08-0.48) | 1.53 (1.35-1.50) |
| **28 Feb 2021** | 50% | 0.31 (0.12-0.59) | 1.56 (1.40-1.70) |
| **15 Jan 2021** | 66% | 0.43 (0.35-0.60) | 1.22 (1.11-1.30) |
| **31 Jan 2021** | 66% | 0.42 (0.35-0.55) | 1.23 (1.13-1.31) |
| **14 Feb 2021** | 66% | 0.49 (0.37-0.67) | 1.23 (1.11-1.32) |
| **28 Feb 2021** | 66% | 0.59 (0.42-0.76) | 1.28 (1.17-1.40) |

**Table S3**

Inferred changes in epidemiological characteristics of B.1.617.2, depending on prior assumptions in the model. Results presented are the median, with the 50% Bayesian Credible Interval, bCI, in brackets. Prior sensitivity analyses assume 50% underreporting in deaths.

| **Sensitivity** | **Prior** | **Inferred epidemiological characteristic** | |
|---|---|---|---|
| | | **Immune escape** | **Transmissibility increase** |
| **Cross-immunity** | Beta(1,1) | 0.43 (0.14-0.79) | 1.38 (1.21-1.58) |
| | Beta(2,1) | 0.22 (0.08-0.48) | 1.53 (1.35-1.50) |
| | Beta(3,1) | 0.12 (0.06-0.26) | 1.60 (1.49-1.67) |
| | Beta(4,1) | 0.10 (0.04-0.18) | 1.62 (1.55-1.68) |
| | Complete escape | 1 | 1.11 (1.08-1.15) |
| | Complete cross-protection | 0 | 1.71 (1.67-1.75) |
| **Timing to 50% waning** | ½ year | 0.57 (0.35-0.76) | 1.51 (1.45-1.58) |
| | 1 year | 0.22 (0.08-0.48) | 1.53 (1.35-1.5) |
| | 2 years | 0.14 (0.08-0.24) | 1.55 (1.45-1.63) |
| **Transmissibility increase** | Gamma(2,2) | 0.13 (0.06-0.29) | 1.60 (1.47-1.67) |
| | Gamma(5,5) | 0.22 (0.08-0.48) | 1.53 (1.35-1.65) |
| | Gamma(10,10) | 0.28 (0.13-0.58) | 1.48 (1.30-1.60) |
| **IFR** | N(0.125,0.02) | 0.73 (0.65-0.81) | 1.11 (1.04-1.17) |
| | N(0.25,0.02) | 0.22 (0.08-0.48) | 1.53 (1.35-1.5) |
| | N(0.5,0.02) | 0.41 (0.20-0.64) | 1.48 (1.39-1.58) |

**Data S1 (separate file)**

GISAID accession IDs, date of collection and lineages of the samples used to calculate lineage proportions in different states from Datasets A and B.

**Data S2 (separate file)**

Cases, Tests, Hospitalizations, ICU Admissions, Deaths data for Delhi from the state level database maintained by NCDC.

**Data S3 (separate file)**

Acknowledgement table for the genomes accessed from GISAID.

**Data S4 (separate file)**

Details of 24 post-vaccination samples

**Data S5 (separate file)**

Raw Ct Values for July 2020-June 2021 for Delhi

**Data S6 (separate file)**

Serosurvey data for the three phases conducted in Delhi (Fig. 2A)

**Data S7 (separate file)**

sVNT assay data Phase I vs Phase II serosuvey (Fig. 2D)

**Data S8 (separate file)**

Data for Subjects with Reinfection (Fig. 2C)

**Data S9 (separate file)**

Data for Subjects without Reinfection (Fig. 2D)

### References


20. F. Wu, S. Zhao, B.Yu *et al.* A new coronavirus associated with human respiratory disease in China. *Nature* **579,** 265-269 (2020).
21. J. Hadfield, C. Megill, S.M. Bell *et al*. Nextstrain: real-time tracking of pathogen evolution. *Bioinformatics* **34,** 4121-4123 (2018).
22. P. Sagulenko *et al.* TreeTime: Maximum-likelihood phylodynamic analysis. *Virus evolution* vol. 4,1 vex042 (2018).
23. B.Q. Minh *et al*. IQ-TREE 2: New Models and Efficient Methods for Phylogenetic Inference in the Genomic Era. *Molecular biology and evolution* **37,** 1530–1534 (2020).
24. G. Yu Using ggtree to Visualize Data on Tree-Like Structures. *Current protocols in bioinformatics* **69**, e96 (2020).
25. D. Wrapp *et al.* Cryo-EM structure of the 2019-nCoV spike in the prefusion conformation. *Science* **367,** 1260-1263 (2020).
26. L. Jun, *et al*. Structure of the SARS-CoV-2 spike receptor-binding domain bound to the ACE2 receptor. *Nature* **581,** 215–220 (2020).
27. S. Fatihi *et al.* A rigorous framework for detecting SARS-CoV-2 spike protein mutational ensemble from genomic and structural features. bioRxiv (2021).
28. E.F. Pettersen *et al*. UCSF ChimeraX: Structure visualization for researchers, educators, and developers. *Protein science* **30,** 70-82 (2021).
29. P. Emsley *et al.* Features and development of Coot. *Acta crystallographica* **66,** 486-501 (2010).
30. S. Flaxman *et al*. Estimating the effects of non-pharmaceutical interventions on COVID-19 in Europe. *Nature* **584**, 257-261(2020).
31. N. Faria *et al.* Genomics and epidemiology of the P.1 SARS-CoV-2 lineage in Manaus, Brazil. *Science* **372,** 815-821 (2021).
32. H.J.T. Unwin *et al*. State-level tracking of COVID-19 in the United States. *Nature communications* **11,1 6189** (2020).
33. W. Feller, On the Integral Equation of Renewal Theory. The Annals of Mathematical Statistics (1941), doi:10.1214/aoms/1177731708.
34. R. Bellman, T. Harris, On Age-Dependent Binary Branching Processes. The Annals of Mathematics (1952), doi:10.2307/1969779
35. N. Brazeau, R. Verity, S. Jenks, H. Fu, C. Whittaker, P. Winskill, I. Dorigatti, P. Walker, S. Riley, R. P. Schnekenberg, others, Report 34: COVID-19 infection fatality ratio: Estimates from seroprevalence (2020) (available at https://www.imperial.ac.uk/mrc-global-infectious-disease-analysis/covid-19/report-34-ifr/)
36. M. Banaji, Estimating COVID-19 infection fatality rate in Mumbai during 2020. medRxiv (2021)
37. S. Purkayastha *et al.* A comparison of five epidemiological models for transmission of SARS-CoV-2 in India. *BMC infectious diseases*. **21,1 533** (2021).
38. M. Pons-Salort, J. John, O. J. Watson, N. F. Brazeau, R. Verity, G. Kang, N. C. Grassly, Reconstructing the COVID-19 epidemic in Delhi, India: Infection attack rate and reporting of deaths. medRxiv (2021).
39. M.V. Murhekar *et al.* Prevalence of SARS-CoV-2 infection in India: Findings from the national serosurvey, May-June 2020. *The Indian journal of medical research* **152,** 48-60 (2020).

40. J. Unnikrishnan *et al.* Estimating under-reporting of COVID-19 cases in Indian states: an approach using a delay-adjusted case fatality ratio. *BMJ open* vol. 11,1 e042584 (2021).
41. R.K. Biswas *et al.* Underreporting COVID-19: the curious case of the Indian subcontinent. *Epidemiology and infection* vol. 148 e207 (2020).
42. J. Hellewell, T. W. Russell, R. Beale, G. Kelly, C. Houlihan, E. Nastouli, A. J. Kucharski, S. Investigators, F. S. Team, C. C. Consortium, others, Estimating the effectiveness of routine asymptomatic PCR testing at different frequencies for the detection of SARS-CoV-2 infections. medRxiv (2020)
43. A. Velumani, C. Nikam, W. Suraweera, S. H. Fu, H. Gelband, P. E. Brown, I. Bogoch, N. Nagelkerke, P. Jha, SARS-CoV-2 seroprevalence in 12 cities of India from july-december 2020. medRxiv (2021).
44. B. Borremans *et al.* Quantifying antibody kinetics and RNA detection during early-phase SARS-CoV-2 infection by time since symptom onset. *eLife* vol. 9 e60122 (2020).
45. J. A. Scott, A. Gandy, S. Mishra, J. Unwin, S. Flaxman, S. Bhatt, Epidemia: Modeling of epidemics using hierarchical Bayesian models (2020), (available at https://imperialcollegelondon.github.io/epidemia/).
46. S. Bhatt, N. Ferguson, S. Flaxman, A. Gandy, S. Mishra, J. A. Scott, Semi-Mechanistic Bayesian Modeling of COVID-19 with Renewal Processes. arXiv (2020).
47. X. Chi *et al*. A neutralizing human antibody binds to the N-terminal domain of the Spike protein of SARS-CoV-2. *Science* **369,** 650-655 (2020).
48. N. Suryadevara *et al*. Neutralizing and protective human monoclonal antibodies recognizing the N-terminal domain of the SARS-CoV-2 spike protein. *Cell* **184,** 2316-2331 (2021).
49. M. McCallum *et al.* SARS-CoV-2 immune evasion by the B. 1.427/B. 1.429 variant of concern. *Science* **373,** 648-654 (2021).
50. Z. Liu *et al.* Identification of SARS-CoV-2 spike mutations that attenuate monoclonal and serum antibody neutralization. *Cell host & microbe* **29,** 477-488 (2021).
51. S.M.C. Gobeil *et al.* D614G mutation alters SARS-CoV-2 spike conformation and enhances protease cleavage at the S1/S2 junction. *Cell reports* **34** (2021).
